# Genomic epidemiology of dengue virus 2 and 3 reveals repeated introductions and exportations of several lineages in Colombia

**DOI:** 10.1101/2025.08.07.25333238

**Authors:** Ricardo Rivero, Vaneza Tique-Salleg, Daniel Echeverri-De la Hoz, Lambodhar Damodaran, Daniela Paternina, Mauricio Santos-Vega, Daniela Torres-Hernández, Diana Davalos, Eduardo López-Medina, Mallery I. Breban, German Arrieta, Jorge Miranda, Verity Hill, Nathan D. Grubaugh, Salim Mattar

## Abstract

Dengue fever, a major mosquito-borne viral disease, is transmitted by *Aedes* mosquitoes and poses a significant global health burden. Despite extensive research, the spatiotemporal dynamics of dengue virus (DENV) lineages in Colombia remain understudied. Here we analyze 11,443 complete genome sequences from Colombia and the Americas to map the genomic epidemiology of DENV-2 and DENV-3. Phylogeographic reconstruction revealed multiple independent introductions and exportations of the DENV-2 II and III lineages, as well as the DENV-3 lineage III C.2, underscoring Colombia’s critical role both as a source and a sink of viral traffic within the Americas. Antigenic profiling demonstrated distinct clustering of emergent lineages in antigenic space, consistent with immune-escape–driven turnover. These results highlight the necessity of sustained, high-resolution genomic surveillance to guide targeted public-health interventions and mitigate dengue transmission across the region.

## INTRODUCTION

Dengue fever is a febrile illness caused by *Orthoflavivirus denguei* (dengue virus, DENV), a member of the family *Flaviviridae*^1^. Global burden estimates indicate 390 million DENV infections occur each year (with 96 million being clinically apparent), and 3.9 billion people live in areas of risk, underscoring the significant public-health impact of the disease^2^. Human infection occurs primarily through the bite of infected mosquitoes belonging to the genus *Aedes*; the geographic range and climatic suitability of *Aedes aegypti* have expanded steadily over the past half-century and are projected to accelerate further under climate-change scenarios^3^. Consequently, almost one-half of the world’s population is currently at risk of dengue infection.

DENV exhibits considerable genetic diversity, characterized by four distinct serotypes (DENV1 to DENV-4), each further subdivided into multiple genotypes^4,5^. Primary infection induces lifelong homotypic immunity but only transient heterotypic protection. Historically, DENV-1 was first isolated as the Mochizuki prototype strain in Japan in 1943 (Genbank: S75335.1), followed by the New Guinea C prototype of DENV-2 in Papua New Guinea in 1944 (Genbank: KM204118.1), H87 (DENV-3) (Genbank: KU050695.1), and H241 (DENV-4) (Genbank: KR011349.2) prototype strains in the Philippines in 1956^6–8^.

In the Americas, transmission has long been dominated by DENV-1 and DENV-2, particularly the II (DENV-2II) and III (DENV-2III) lineages. The 2023–2025 continental epidemic was unprecedented: by the end of 2024 the Region of the Americas had reported 13 million suspected dengue cases—more than triple the previous regional record^9^. Brazil alone accounted for about 6.6 million cases during 2024, while Colombia and Ecuador reported 321,907 and 57,712 cases, respectively, during 2024^9–11^. The same ecological window has coincided with a resurgence in Yellow Fever cases—104 cases between 2024 and May 30th, 2025—highlighting that the climatic conditions driving dengue could favor the circulation of other arboviruses in the region. Climate anomalies associated with the 2023–2024 El Niñepisode substantially increased vector suitability and amplified year-round transmission across equatorial South America^12^.

Previous phylogeographic studies have documented frequent inter- and intra-continental movement of DENV lineages. Endemic circulation in Colombia, Puerto Rico, and Brazil is sustained by the cryptic co-circulation of several serotypes and continual lineage importation from neighboring regions and beyond^13–18^. Despite these insights, high-resolution genomic surveillance remains essential to disentangle the drivers of current outbreaks. Applying the newly proposed dengue lineage-assignment system, we show that lineages 2II F.1.1.2 and 2III D.2 dominate recent DENV-2 transmission in the Americas. Our phylogeographic reconstruction highlights Colombia’s pivotal role as a bridge for viral dissemination between South America, the Caribbean, and North America. We also report that undetected introductions from Venezuela, as well as cryptic transmission, could have driven the circulation of DENV-3III C.2 in Colombia during 2024 and 2025, but went largely undetected due to the continuous decrease in the sequencing effort of DENV cases in Venezuela since 2006. We also found a significant antigenic divergence between the two competing DENV-2 lineages (2II F.1.1.2 and 2III D.2), which suggests that the replacement dynamics observed after the invasion of 2II F.1.1.2 might have been driven by antigenic escape to homotypic immunity, highlighting the role of genetic diversity in the population susceptibility to DENV. Together, our findings highlight the importance of sustained genomic and epidemiological surveillance and their critical role in detecting introduction events, characterizing cryptic transmission, and antigenic variation to inform timely public-health responses across the region.

## RESULTS

### Multiple DENV serotypes and lineages have circulated in Colombia between 2018 and 2024

Since 2018, the circulation of DENV in Colombia has been extensively dominated by DENV-1 genotype V. While the number of DENV cases decreased across the five Colombian regions during the SARS-CoV-2 pandemic, 2021 was characterized by a sharp increase in the proportion of DENV cases associated with the 2III D.2 lineage, which rose to 95% during 2021. By 2022, 2III D.2 was quickly replaced by 2II F.1.1.2, continuing to dominate the transmission of the virus since 2022 (Fig. 1A). This genotype has been reported to be introduced into the Americas from Southeast Asia in 2017^19^, and quickly established sustained regional transmission. Once established in the region, this lineage has continued to evolve, accumulating mutations of concern such as NS4B V91A that confers resistance to Mosnodenvir, while quickly rising to a majority share in the French Caribbean islands^20^. Between 2022 and 2024, multiple DENV serotypes and genotypes have been transmitted in Colombia, with the notable resurgence of 3III C.1 and C.2, which had not been detected in Colombia since the end of 2017, reaching 34% in August of 2023(Fig. 1A). To further describe the epidemiological situation of Colombia during the latest DENV epidemic, we subdivided the country into its 5 regions: Amazonian, Andean, Caribbean, Orinoquia, and Pacific (Fig. 1B). Then, we calculated the mosquito suitability index (index P)^21^, and assessed its association with weekly dengue case counts resulting in a significant correlation between the observed increase in index P and dengue incidence across all regions, with the strongest associations observed in the Caribbean (*ρ* = 0.38, *p <* 0.00001) and Orinoquia (*ρ* = 0.38, *p <* 0.00001) regions (Fig. 1C, Fig. S3). Lagged correlation analysis further indicated that increases in index P preceded rises in case counts by 2–6 weeks. A particular case was the Amazonian region where a lead of 6 weeks yielded a strong inverse association (*ρ* = –0.53, *p <* 10^−11^), showing a decoupling between vector suitability and human cases that could be attributed to under reporting of cases. In 2024, which reported an unprecedented 321,907 DENV cases, these trends coincided with 2024 being an El Niñyear^22^, which likely facilitated the reproduction and life-cycle of *Aedes aegypti* and *Aedes albopictus*, which are the two main vectors of DENV in Colombia^23,24^.

**Figure 1.**
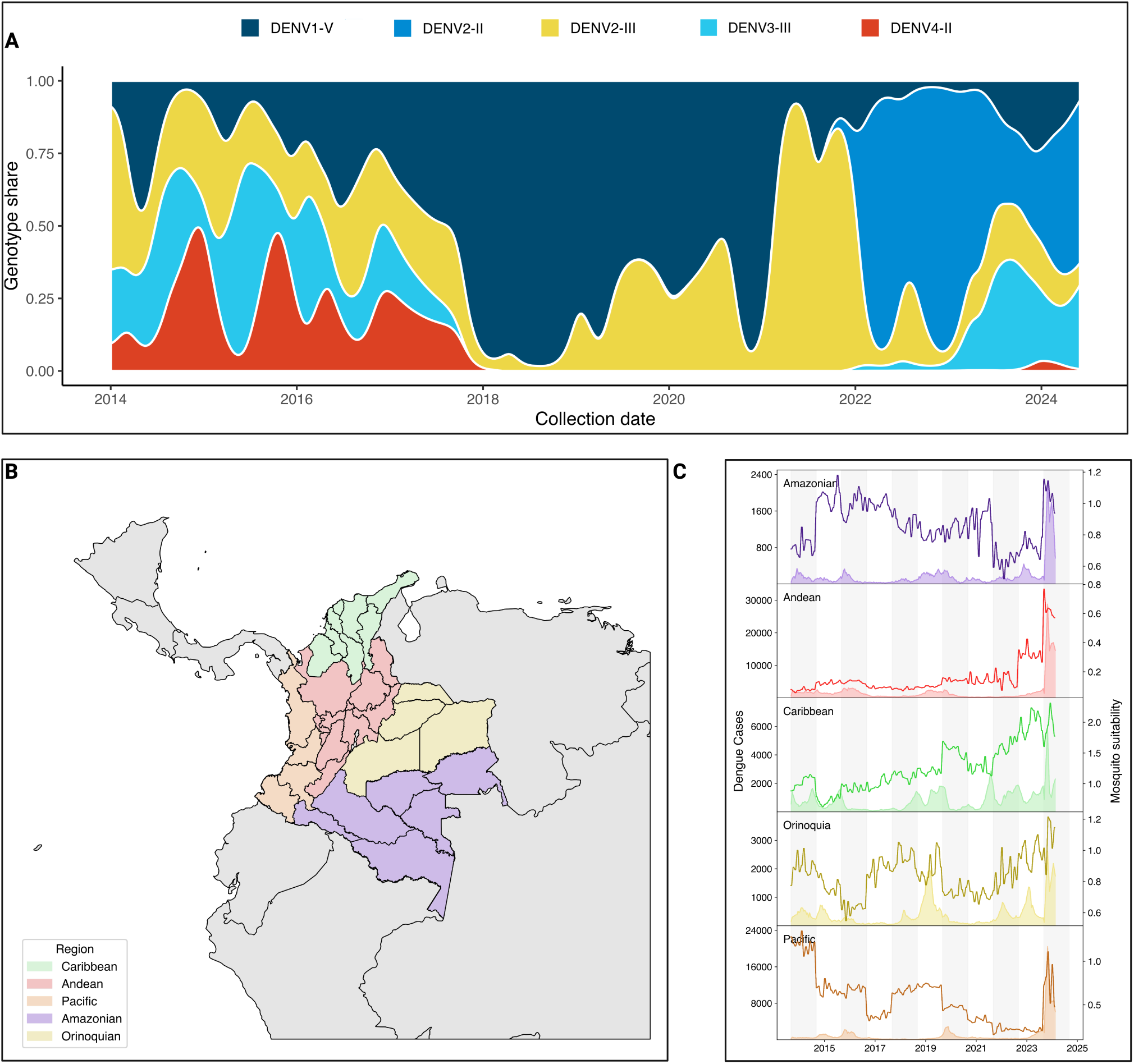
Genomic and epidemiological characteristics of DENV circulation in Colombia. A) Smoothed stacked proportion of DENV lineage circulation in Colombia between 2014 and 2024, data gathered from EpiArbo and transformed into timeseries of lineage prevalence based on the assigned clades according to dengue-lineages aggregated into genotypes. B) Map of the five natural regions of Colombia subdivided into its corresponding administrative units, referred as ”departments”. C) Region-aggregated time series of DENV cases (area shaded timeseries, left-hand axis) and Mosquito suitability index (index P) as calculated from metereological data published by IDEAM using the MVSE package.

Collectively, these observations suggest that the unprecedented 2024 dengue epidemic in Colombia was in part propelled by (i) successive serotype/genotype replacement events culminating in the establishment of the II DENV-2 lineage, and (ii) climatic conditions that amplified *Aedes* population fitness and virus transmission potential.

### The circulation of the Colombian DENV-2III D.2 lineage was characterized by cryptic transmission and repeated exportation events

Our analysis identified a substantial circulation of lineage 2III D.2, comprising a total of 316 sequences. The time to the most recent common ancestor (tMRCA) for this lineage was estimated as December 22, 2004 (95% HPD: February 26, 2003, to December 31, 2005). Phylogenetically, 2III D.2 derives from from the broader III genotype first introduced into Colombia during the 1970s^25^. Within Colombia, lineage 2III D.2 experienced prolonged cryptic transmission from 2010 to 2021, with sporadic detections occurring in 2015 and 2016. The lineage re-emerged prominently in 2021, subsequently co-circulating alongside other DENV-2 and DENV-3 lineages during the significant dengue outbreak from 2022 to 2024. During this period, the lineage exhibited an extended median trunk reward time of 147.27 years (95% HPD: 103.98–182.45), that is, the cumulative branch length associated with 2III D.2 at a given state (Colombia in this case) across the posterior distribution of trees (Fig. 2A)^26^.

**Figure 2.**
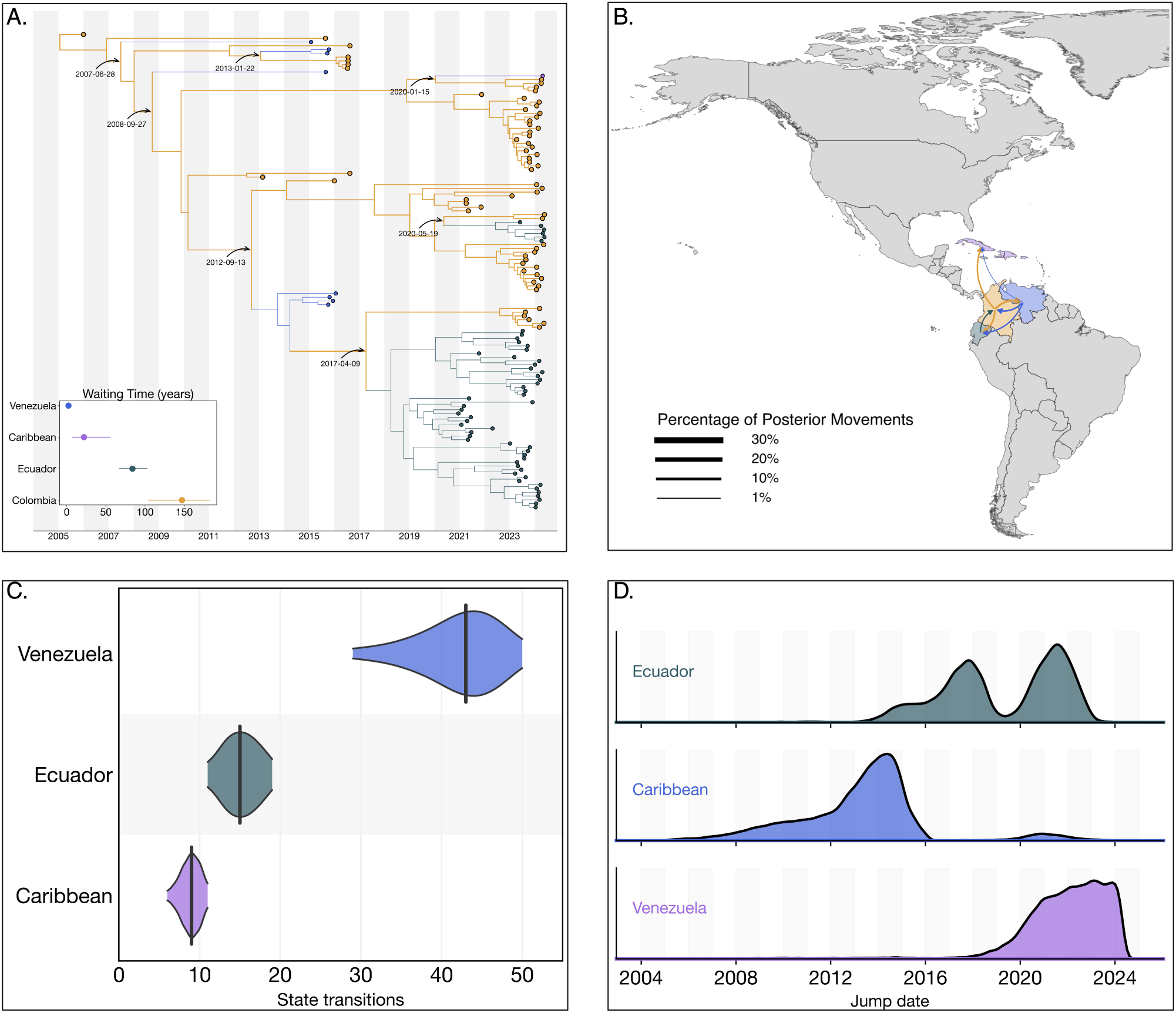
Phylogeographic reconstruction of the DENV-2III D.2. A) Maximum clade credibility (MCC) tree of the inferred phylogeography of DENV-2III.D.2 in the Americas, introduction events are highlighted using curved arrows, with median tMRCAs annotated as dates. Median trunk reward time per country and 95% HPD are displayed in the inset plot at the lower-left of the panel. B) Map representing the percentage of movements (Markov jumps) between countries, transitions representing ¡1% of the total movements were not plotted, and the line thickness represents the percentage of movements between country pairs. Countries were colored to differentiate them in the map, and the arrow is colored according to the movement’s country of origin. C) Violin plot representing the median and 95% HPD of the number of state transitions from Colombia into other countries as reconstructed from the posterior set of trees. D) Density distribution of the inferred timing of viral lineage exportations from Colombia into Ecuador, the Caribbean and Venezuela.

Phylogeographic reconstructions revealed limited exportation events from Colombia to other countries, notably involving repeated exports into the Caribbean, Ecuador and Venezuela between 2007 and 2013. These exports resulted in transient clusters within Venezuela and isolated single-case detections (singletons) in the Caribbean by 2020. More recently, independent exports of lineage 2III D.2 from Colombia into Ecuador occurred on April 09, 2017, and May 19, 2020, subsequently giving rise to substantial transmission clusters within Ecuador.

A complete reconstruction of viral lineage movements indicated a minority (3,189/78,273, 4.07%) of re-introduction events from Ecuador back into Colombia (Fig. 2B). These events, inferred across the full posterior (BF = 3.66), were not captured by the maximum clade credibility (MCC) tree, likely due to insufficient sampling density. In contrast, summarizing discrete state transitions at internal nodes across 10,000 posterior trees yielded a median of 43 introductions into Venezuela, 9 into the Caribbean, and 15 into Ecuador from Colombia (Fig. 2C). Temporal density estimates from the full posterior distribution suggest that exports from Colombia into the Caribbean most likely occurred between 2008 and 2016, well before the 2020 introduction date inferred by the MCC tree and the first sampling date in 2024 (Fig. 2D). These introductions are strongly supported with a Bayes Factor of 28.01 and a posterior probability of 0.926, indicating high confidence in the existence of one or more transmission events during this period, likely reflecting the role of asymptomatic or undiagnosed travelers returning from Colombia^27^.

Collectively, these results highlight the critical role of cryptic transmission in facilitating the resurgence of lineage 2III D.2 within Colombia during the 2022–2024 dengue outbreak and underscore Colombia’s significance as a source for regional dissemination of the lineage across neighboring countries in the Americas. With a strong support for exports from Colombia to the Caribbean (BF = 28.01), Ecuador (BF = 21.49), and Venezuela (BF = 17.66), consistent with Colombia acting as a major hub of regional viral spread.

### Extensive spread of DENV-2II F.1.1.2 across the Americas and establishment of domestic clades in Colombia

In 2022, lineage 2III D.2 was displaced by lineage 2II F.1.1.2. The median time to the most recent common ancestor (tMRCA) of lineage 2II F.1.1.2 was estimated to be October 02, 2017 (95% HPD interval: June 26, 2008, to February 17, 2019), corresponding to its introduction from Bangladesh into Peru. Phylogeographic reconstruction demonstrated extensive dissemination of this lineage throughout the Americas, with significant viral exchange events identified among Brazil, Bolivia, Colombia, Ecuador, Peru, Paraguay, and the United States (Fig. 3A).

**Figure 3.**
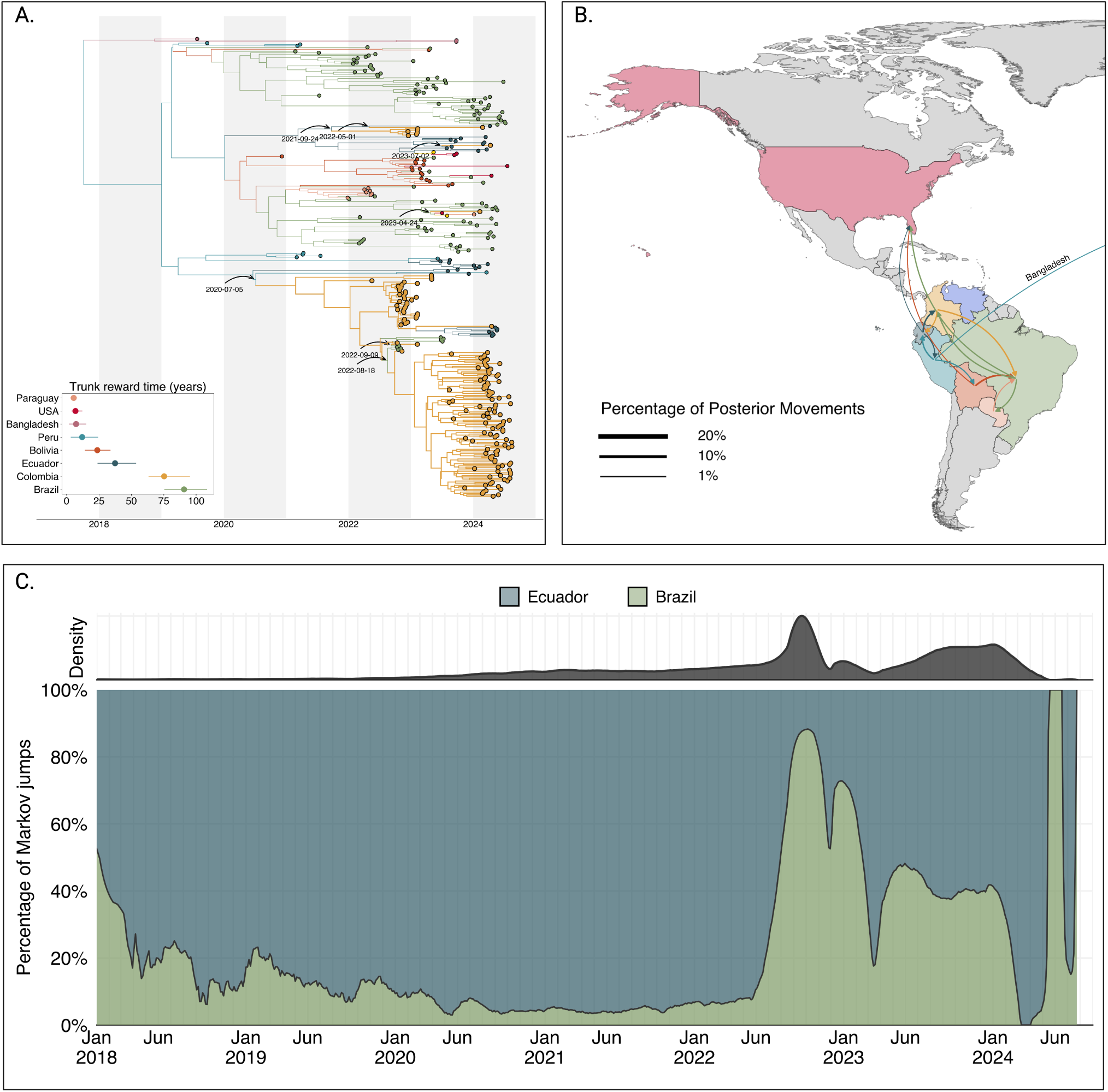
Phylogeographic reconstruction of the DENV-2II F.1.1.2. A) Maximum clade credibility (MCC) tree of the inferred phylogeography of DENV-2II.F.1.1.2 in the Americas, introduction events are highlighted using curved arrows, with median tMRCAs annotated as dates. Median trunk reward time per country and 95% HPD are displayed in the inset plot at the lower-left of the panel. B) Map representing the percentage of movements (Markov jumps) between countries, transitions representing ¡1% of the total movements were not plotted, and the line thickness represents the percentage of movements between country pairs. Countries were colored to differentiate them in the map, and the arrow is colored according to the movement’s country of origin. A pair of transitions between Peru and Bangladesh is represented in the map as the reconstructed location at the root of 2II F.1.1.2 in the Americas, Bangladesh is not shown in the map. C) Stacked area plot of the percentage of introductions into Colombia over time, at the top of the panel, the inset shows the density distribution of introduction count over time.

Our analyses revealed five independent introductions of lineage 2II F.1.1.2 into Colombia. The earliest introduction occurred from Ecuador on July 05, 2020, followed by a period of cryptic transmission prior to the establishment of two substantial domestic clusters in 2023 and 2024. Additional independent introductions occurred during 2021, 2022, and 2023, resulting in smaller clusters and isolated singletons. Furthermore, our analysis indicated sustained and extensive circulation of lineage 2II F.1.1.2 within Colombia, Ecuador, and Brazil, with median trunk reward times estimated at 90.69, 75.30, and 37.44 years, respectively (Fig. 3A).

To elucidate viral migration patterns more explicitly, we quantified viral lineage movements using Markov jump analyses. This analysis indicated frequent and substantial viral transitions involving Colombia, suggesting that Colombia might have a central role as a bridging region for lineage 2II F.1.1.2 between South and North America (Fig. 3B). Of all well-supported transitions (*>* 1% of total movements), 12.78% originated from Colombia and 17.69% were directed into Colombia, underscoring its role as both a major source and recipient of regional viral movement (Fig. 3B,C). To evaluate this hypothesis, we reconstructed the complete history of viral transitions using the maximum clade credibility tree (MCC) and implemented a Poisson Generalized Linear Model (GLM). The GLM quantified the impact of excluding Colombia as a source or sink of viral introductions while incorporating time as a categorical variable to control for temporal variations in transmission intensity.

The results of this analysis demonstrated that, when controlling for time and directionality of movement, removal of Colombia significantly reduced the log count of Markov jumps by 0.2258, corresponding to a substantial reduction of approximately 21.1%. These findings statistically confirm that Colombia functioned as a critical node that facilitates transmission of the 2II F.1.1.2 lineage between the southern and northern American regions.

To further test whether this apparent centrality could reflect sampling bias, we performed a complementary bootstrap-based source-sink score (SSS) analysis based on the method proposed by Lyu et al^28^, which quantifies the net directional imbalance of viral flow (positive = source-leaning; negative = sink-leaning). We estimated SSS values under two independent phylogeographic frameworks: (i) the full maximum-likelihood tree of 1,795 DENV2-II F.1.1.2 sequences without subsampling and (ii) the time-scaled BEAST MCC tree based on the temporally and geographically subsampled dataset. Three bootstrap strategies (event resampling, time-stratified resampling, and tip jackknifing) were each iterated 5,000 times to obtain 95% confidence intervals.

Under the ML framework, Colombia’s mean SSS was 0.375 (95%CI:[–0.13, 0.75]) across bootstraps, indicating a mild source-leaning balance. Under the BEAST reconstruction, the mean SSS was –0.17 (95%CI:[–0.67, 0.33]), a slightly sink-leaning but near-neutral tendency. Both confidence intervals overlapped zero, and the magnitudes were modest (*|SSS| <* 0.4), consistent with a hub-like role in which Colombia mediates bidirectional north–south viral exchange rather than acting as a pure source or sink (Fig. S5). These results reinforce the Markov-jump findings while demonstrating that the inferred centrality of Colombia remains robust to differences in phylogenetic inference and to bootstrapped resampling of transition events.

### Reciprocal introduction dynamics between Colombia and Venezuela drove the emergence and maintenance of DENV-3III C.2

The 2023–2024 dengue epidemic in Colombia was also marked by the reemergence of lineage C.2 of dengue virus serotype 3 genotype III (DENV-3III C.2). Our analysis revealed reciprocal introduction dynamics primarily between Colombia and Venezuela, with minimal evidence of lineage circulation detected elsewhere (only two sequences identified in other regions). Phylogeographic reconstruction identified multiple introductions of lineage 3III C.2 from Venezuela into Colombia occurring between 2000 and 2007, subsequently leading to small, transient clusters of infections within Colombia from 2014 to 2017 (Fig. 4A).

**Figure 4.**
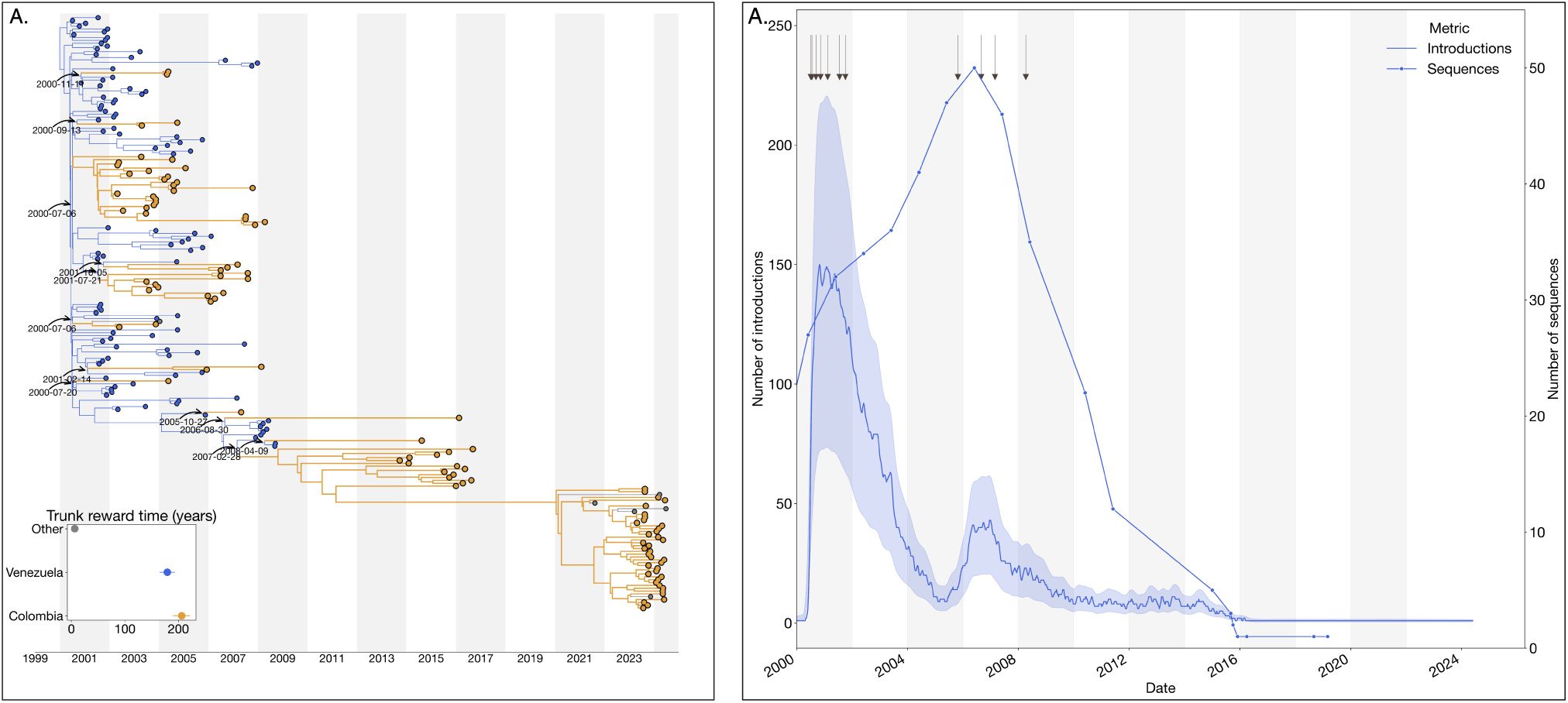
Phylogeographic reconstruction of the DENV-3III C.1. A) Maximum clade credibility (MCC) tree of the inferred phylogeography of DENV-3III C.1 in the Americas, introduction events are highlighted using curved arrows, with median tMRCAs annotated as dates. Median trunk reward time per country and 95% HPD are displayed in the inset plot at the lower-left of the panel, countries with *<* 3 sequences were grouped into ”Others”. B) Time series of the reconstructed number of introductions (median + 95% HPD) and number of DENV sequences generated in Venezuela over time (line with dots). The arrows at the top of the figure represent the time of the inferred introductions in the MCC.

Additionally, evidence of cryptic transmission starting around 2011 preceded the prominent reemergence of lineage 3III C.2 in 2023. To evaluate the hypothesis that this cryptic transmission reflected the reduced genomic surveillance and sequencing activities in Venezuela rather than changes in epidemiological factors such as human mobility or cross-border connectivity, We reconstructed the complete jump history and conducted a Spearman correlation analysis, demonstrating a significant positive association between the annual decline in available Venezuelan viral genomes and the number of introductions detected (*ρ* = 0.67, p = 0.002) (Fig. 4B). These findings support the notion that persistent introduction events may have gone undetected due to reduced sampling efforts in Venezuela, emphasizing the importance of sustained genomic surveillance for accurate tracking and response to dengue transmission dynamics.

### DENV-2II F.1.1.2, DENV-2III D.2 and DENV-3III C lineages displaced earlier strains by occupying novel antigenic clusters

The Colombian dengue epidemics of 2019–2024 were characterised by two successive lineage sweeps in DENV-2 and the re-emergence of DENV-3 genotype III (Fig. 1A). To determine whether these turnovers reflect immune-driven selection rather than purely demographic processes, we integrated (i) structure-mapped conservation scores for the envelope (E) homodimer (Fig. 5A), (ii) an antigenic two-dimensional embedding of Colombian E-protein sequences, and (iii) residue-level Shannon-entropy profiles (Fig. 5). To visualize these differences, we used a t-SNE projection of pairwise antigenic distances between the Envelope protein sequences, which places each major lineage in separate clusters. DENV-2III D.2, which was dominant in 2019–2021, is clearly separated from the replacement lineage DENV-2II F.1.1.2, indicating minimal antigenic cross-recognition. Likewise, the recently resurgent DENV-3III C.1/III C.2 clusters (greens/browns) form an isolated neighborhood (Fig. 5B). These clusters reflect the divergence observed in the unrooted phylogeny between the III and II genotypes. These abrupt change in the t-SNE coordinates of the viral envelopes is consistent with selective sweeps rather than gradual drift.

**Figure 5.**
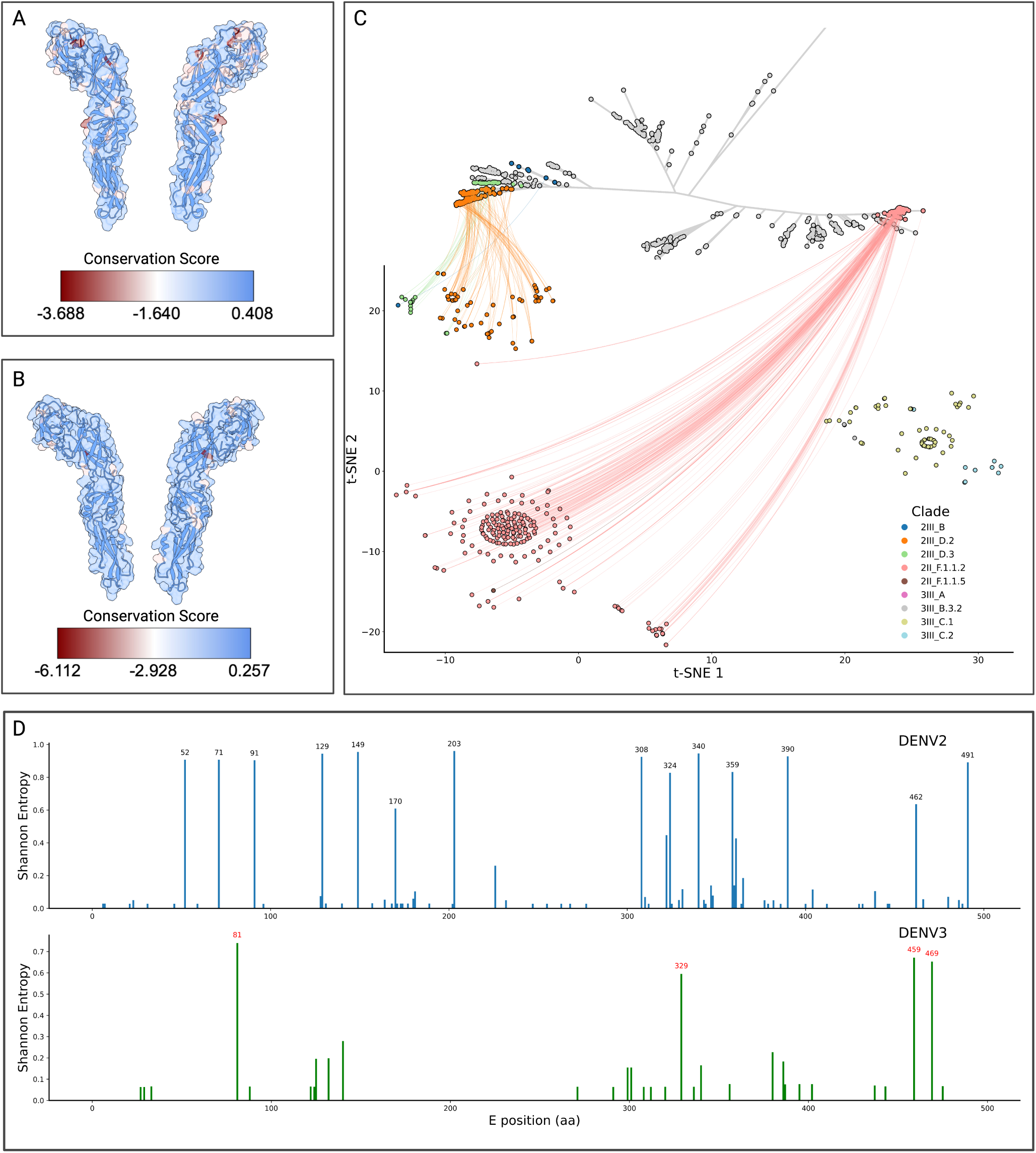
Structural and antigenic variability of DENV-2 and DENV-3 in Colombia. A) Exploded view of the envelope of DENV-2 (PDB: 4UTC), with superimposed surface and ribbon cartoon view and coloring based on their conservation score as determined from an alignment of all DENV-2 envelope sequences sampled in Colombia. B) Exploded view of the envelope of DENV-3 (PDB: 1UZG), as described for panel A. C) Unrooted maximum-likelihood phylogenetic tree of the global diversity of DENV-2 with Colombian sequences placed onto the global backbone and colored by clade. The leaves of the tree are connected to their corresponding t-SNE representation, which was performed to aggregate the envelope sequences according to their sequence-level differences. A cluster of DENV-3 sequences was added to visualize its clustering with DENV-2 sequences, but is not included in the unrooted tree. D) Positionwise Shannon entropy (diversity) of the envelope of DENV-2 and DENV-3 lineages sampled in Colombia, those protein positions with a Shannon entropy > 0.6 are labeled in black and red for DENV-2 and DENV-3, respectively.

We then mapped conservation values onto prefusion E-homodimer crystal models, revealing discrete surface patches of high variability in both serotypes (Fig. 5D). Residue-resolved entropy scans pinpointed the same sites—e.g. positions 52, 71, 91, 129, 149, 170, 203, 308, 324, 340 359, 390, 462, and 491 in DENV-2; 81, 329, 459, and 469 in DENV-3—as being the most variable compared to the reference (Fig. 5D). Interestingly, these residues localize in the fusion loop, lateral ridge, and domain III dimer interface (Fig. 5A,B), which are targeted by strongly neutralizing antibodies^29,30^. The perfect concordance between structural variability and sequence entropy argues that each emergent lineage acquired mutations at key neutralizing epitopes, thereby evading pre-existing population immunity. Taken together, the data show that (i) each victorious lineage occupies a previously unfilled region of antigenic space, (ii) distinguishing substitutions are confined to well-defined neutralizing epitopes, and (iii) those substitutions arose just prior to or during the epidemiological expansion of the lineage. These observations support a model in which immune-escape variants establishing distinct antigenic clusters out-compete resident strains, driving the rapid replacements observed in 2022 (DENV-2) and 2023–2024 (DENV-3). This mechanism parallels the punctuated antigenic shifts seen in influenza and highlights the importance of real-time genomic surveillance to anticipate vaccine mismatch and antiviral resistance.

## DISCUSSION

The results of the present study reveal the complex circulation dynamics of the DENV-2III D.2 lineage in Colombia. The analysis indicates that this lineage was established in Colombia around 2004. These support the hypothesis that the lineage descends from the III line introduced in Colombia in the 1970s^17,31^. Subsequently, the 2III D.2 lineage showed a cryptic transmission pattern between 2010 and 2021 before generating the epidemic outbreaks of 2022-2024. The analyses also suggest a successful adaptation of the lineage to local conditions, possibly mediated by biological and ecological factors. In addition, its circulation in unmonitored or sylvatic reservoirs or populations with waning immunity could explain their maintenance during interepidemic periods. Furthermore, reemergence in 2021 and co-circulation with DENV-2 and DENV-3 during the 2022–2024 outbreak indicate that these lineages may expand rapidly. This pattern exposes the difficulties in the early detection of outbreaks when there is asymptomatic or underdiagnosed transmission^13^.

Although surveillance gaps provide a parsimonious explanation for the apparent cryptic transmission of DENV-2III D.2, we acknowledge that phenotypic variation among closely related viruses could also modulate lineage persistence and epidemic potential. Experimental work has shown that even limited amino acid substitutions can produce marked changes in DENV-2 replicative fitness in both human and Aedes aegypti cells^32,33^. However, sub-surveillance transmission of low-frequency lineages is a well-documented phenomenon in dengue and other arboviruses, where genomic sampling preferentially captures the most prevalent variants at any given time. Phylogenetic reconstructions can thus reveal previously undetected local circulation that occurred below the detection threshold but later seeded resurgence. While genetic or phenotypic differences remain plausible contributors, the most likely scenario consistent with our data is localized persistence under reduced detection rather than the emergence of a phenotypically distinct sublineage.

A relevant finding is the role of Colombia as a central point in the regional dispersal of the 2III D.2 lineage. Although viral flow from Colombia to other countries is limited, phylogeography analysis demonstrates multiple dissemination events with establishment patterns in recipient countries. In Ecuador, the 2III D.2 lineage managed to establish itself and generate sustained transmission. Possibly favored by terrestrial connectivity with Colombia and similarity with vector ecosystems. Meanwhile, in the United States and Venezuela, the impact was limited due to the inefficient maintenance of the virus generated by population or ecological factors or by the scarce genomic surveillance for the latter reflection variations in surveillance capabilities. Likewise, the reintroduction from Ecuador to Colombia, possibly facilitated by asymptomatic travelers, suggests a bidirectional flow of the virus that could underscore the complexity of transmission routes. This highlights the importance of border regions as epidemiological monitoring points to implement robust monitoring strategies, since even if viremic travelers are frequently asymptomatic^27^, random or sentinel genomic screening at travel hubs can still provide early information about viral traffic between countries, complementing established community-based surveillance rather than preventing transmission.

Several demographic and geographic features plausibly explain Colombia’s role as a regional transmission node for dengue. First, travel-based genomic surveillance has shown that viral movement across the Americas can be effectively traced through infected travelers, with repeated detections along major air and overland corridors^27^. Second, Colombia has experienced record international mobility in recent years: international visitor arrivals reached historic highs, with 5.86 million non-resident visitors in 2023, reflecting dense air connectivity across Bogotá, Medellín, Cali, and Cartagena^34^. Third, sustained cross-border population flows—especially the large Venezuelan diaspora residing in or transiting through Colombia—create persistent person-movement links across the Caribbean–Andean axis^15,35^. Together with favorable *Aedes* ecologies across altitudinal gradients, these mobility networks help explain Colombia’s hub-like pattern in our phylogeographic reconstructions (near-neutral SSS; modest, overlapping CIs), without requiring a strictly unidirectional source–sink interpretation.

The evolutionary dynamics of DENV-2II F.1.1.2) and DENV-3III C.2) reveal convergent epidemiological patterns, establishing Colombia as a pivotal hotspot for dengue transmission in the Americas. Phylodynamic analyzes demonstrate that both serotypes underwent multiple crossborder introductions followed by sustained local transmission, culminating in their emergence as dominant lineages during the 2023-2024 epidemics. In particular, Bayesian phylogeographic reconstruction and discrete-state Markov jump models position Colombia as an epidemiological conduit between South and North America. The extended persistence of DENV-2II F.1.1.2 in Colombia (90.69 years; 95% HPD: 85.4-96.1) contrasts sharply with the rapid lineage turnover observed in Brazil (37.44 years), suggesting viral adaptation to local ecologies, possibly facilitated by adaptive mutations in NS5 non-structural protein substitutions analogous to those reported in Nicaraguan strains^36^

Consistently, the re-emergence of DENV-3III C.2 was associated with introductions from Venezuela (2000-2007) followed by cryptic transmission since 2011. Although limited detection outside the Colombian-Venezuelan region indicates focused geographic dispersion, a critical finding is the significant positive correlation (*ρ* = 0.67, p = 0.002) between the decrease in sequenced genomes in Venezuela and the number of introductions detected in Colombia, revealing that the reduction in Venezuelan genomic surveillance masks continuous viral flows. Recent studies show that the geographical proximity between Colombia and Venezuela facilitates the bidirectional exchange of various dengue serotypes and genotypes, as observed in the 2015-2016 outbreak in the border region, where frequent introductions of DENV-1 and DENV-2 were identified from Venezuela to Colombian departments such as Norte de Santander^37^. However, irregular surveillance distorts the perception of viral dynamics, reflecting sampling gaps and not the absence of virus transmission, which has also been reported for ZIKV and CHIKV^15,27,38,39^.

The observed dynamics in Colombia show similarities with previous studies in other regions of the country^13^ (Martínez et al., 2024). The pattern of multiple introductions followed by local establishment of DENV-2 resembles that documented by Thongsripong et al. (2023) for the NI3B.3 lineage in Nicaragua (2III D.1.2), while the cryptic transmission of 3III C.2 parallels findings from Angola due to surveillance limitations^40^. However, the transmission dynamics in Colombia are complex. Colombia acts as a recipient of introductions from Ecuador and Venezuela, and as a source of regional dispersion, which exceeds that observed in Brazil^41^. The underestimation of DENV-3 introductions into Colombia reflects the global geographic sampling biases identified by Wei and Li (2017), demonstrating how diagnostic deficiencies can create surveillance blind spots to track disease dynamics^42^.

The epidemic reemergence of both lineages in 2023-2024 raises questions about possible immunological factors involved. The context of hyperendemicity in Colombia, with concurrent circulation of multiple serotypes of DENV and other arboviruses (Zika, Chikungunya and yellow fever virus), could have generated conditions analogous to those documented in post-Zika Nicaragua^36^ and Angola^40^, where cross-immunity would modulate viral severity and persistence. In particular, the temporal coincidence between DENV-3III C.2 and the circulation of 2II F.1.1.2 from DENV-2 suggests possible serological interactions between serotypes. Antigenic analyses reveal that such events reflect a pattern of selection driven by population immunity. The projection of t-SNE of the antigenic distances of the E protein showed a clear separation between the dominant lineage of DENV-2III D.2 in 2019–2021 and its replacement (DENV-2II F.1.1.2), along with an isolated grouping of the reemerging lineages of DENV-3III C.1 and DENV-3III C.2), could indicate a limited cross-recognition by pre-existing antibodies.

Structural examination of protein E homodimers identified regions of high variability (fusion loop, lateral crest, domain III dimeric interface). The key sites in DENV-2 and DENV-3 exhibited significantly elevated Shannon entropy against reference strains, evidencing selective pressure. Structural variability, sequence entropy, and the epidemiological event of substitutions reveal that emerging lineages acquire mutations in immunodominant epitopes to evade population immunity. These findings align with the global evolutionary dynamics of dengue. Studies in Thailand showed that serotypes evolve with periodic fluctuations in antigenic similarity linked to epidemic magnitude, with an evasion of homotypic immunity observed^43^. Similarly, recurrent fluctuations in E gene divergence were documented in India^44^, where the emergence of the DENV-4-Id lineage with convergent drift towards DENV-1/DENV-3 suggests that antibody-dependent enhancement could shape the evolution towards variants with heterologous subneutralizing responses.

While our analyses emphasize variability within the envelope (E) gene because of its central role in antibody recognition, we acknowledge that adaptive differences could also arise outside of the structural proteins. Regulatory and untranslated regions have been shown to affect DENV replication and transmission efficiency through effects on RNA folding, translation, and modulate fitness independently of antigenicity. Therefore, the patterns of antigenic clustering observed here should be interpreted as consistent with—but not direct evidence for—immune-driven selection. Comprehensive analyses integrating whole-genome variability, including the 5^′^*/*3^′^ UTRs and experimental neutralization data, will be essential to test these hypotheses.

In Colombia, the formation of ’antigenic clusters’ underscores the central role of immune selection in viral replacements. Such antigenic plasticity, mediated by strategic mutations in the E protein, has critical implications for control. The close epidemiological relationship between Colombia and neighboring countries highlights the need to implement coordinated regional genomic surveillance systems. These findings emphasize the need to include viral mobility patterns and local factors that modulate the emergence and persistence of specific lineages in epidemiological surveillance of the disease. Combining advanced genomic tools with spatially explicit epidemiological models could improve the ability to predict future outbreaks^40,41^. In particular, the scenario between Colombia and Venezuela warns about how the asymmetry in the surveillance system can generate critical points with consequences for epidemics at border departments.

### Limitations of the study

In this study, we discuss the role of preexisting immunity, epidemiological, and environmental variables in the emergence, establishment and circulation of novel dengue lineages in the record-breaking number of cases between 2018 and 2024. However, further analyses are required to test the extent of cross-protective immunity’s role in shaping the epidemiology of dengue in Colombia. Despite previous studies validating the utility of sequence and structure based approaches as surrogates of antigenic cartographies^45^, antibody-neutralization studies remain the gold standard in quantifying the risk of infection and severe disease upon secondary infection. Additionally, we acknowledge that phylogenetic and phylogeographic inferences can be affected by sampling bias. Differential sequencing intensity—both within Colombia and across neighboring countries such as Venezuela and Ecuador—likely impacts estimates of lineage longevity, timing of introduction, and inferred directionality of viral movement. We also acknowledge that a nation-wide and sustained sequencing effort is essential for continuing to understand the finegrained evolutionary dynamics of dengue in Colombia and its effects in terms of vaccine efficacy, and effectiveness of public health interventions. Finally, we did not explicitly model human mobility (e.g., airline passenger volumes, road traffic, or migration flows); integrating such covariates would enable a more mechanistic assessment of how movement patterns contribute to the observed hub-like role of Colombia^27^.

## STAR METHODS

### Resource availability

#### Lead contact

Requests for further information and resources should be directed to and will be fulfilled by the lead contact, Ricardo Rivero.

#### Data and code availability

- All the generated sequences have been deposited and published in GenBank under the accession numbers 1425-1490.
- The findings of this study are partially based on metadata and genome sequences associated with 26,633 DENV-2 and 10,753 DENV-3 sequences available on GISAID up to July 26, 2025, and accessible at https://doi.org/10.55876/gis8.250726uw and https://doi.org/10.55876/gis8.250726uh.
- All code required to reproduce the figures and analyses done in this study is available at Github (https://github.com/RicardoRH96/DENV_Epi).
- Any additional information required to reanalyze the data reported in this work paper is available from the lead contact upon request.

### Method details

#### Sample collection and whole-genome sequencing

Serum samples were collected from patients who received care due to virologically confirmed dengue (VCD) at Hospital Universitario del Valle (HUV), Hospital San Jerónimo de Monteria (HSJM) and Clinica Salud Social (CSS) in the cities of Cali, Montería and Sincelejo during the 2023-2024 dengue season (15 April 2023 until 30 Aug 2024). Suspected dengue was defined as episodes in which a diagnostic dengue test was performed at HUV based on clinical suspicion. VCD was diagnosed in patients with a documented fever (¿38 °C) of less than 7 days duration and one of the following manifestations: Headache, retroocular pain, myalgia, arthralgia, nausea, vomiting or rash, (12) along with a positive NS1 for patients with symptom onset of 5 days or less, or IgM dengue enzyme-linked immunosorbent assay (ELISA) for patients with symptom onset of 6 days or more. DENV evaluation was performed with the VIDAS® dengue panel (bioMérieux, Marcy-l’Étoile, France). For each collected serum sample, 140 µL was used for viral RNA extractions using the QIAamp Viral RNA Mini Kit (QIAGEN Inc., Germantown, MD, USA.) according to manufacturer’s instructions. Then, identification of DENV serotypes (DENV-1 to -4) was performed on all samples using the CDC DENV-1-4 rRT-PCR Multiplex Assay for DENV typing^46,47^. Whole-genome sequencing was performed using DengueSeq^48^. Bioinformatics analysis, including primer trimming and consensus generation, was conducted with the iVar pipeline^48^, althought partial UTRs were sequenced, these were masked from the analyses to preserve the robustness of the phylogenetic reconstruction. Samples with *≥*5% genome completeness were assigned DENV lineages using Nextclade^5^. The lineage classifications were used to verify the PCR-based serotype calls. DENV genomes greater than 70% completeness are available at National Center for Biotechnology Information BioProject (https://www.ncbi.nlm.nih.gov/bioproject; accession no. PRJNA1132139) and accession no. OQ813507. Samples from HSJM can be accessed at Epicov.org under the accession numbers EPI ISL 17976304 and EPI ISL 17977444.

#### Collection of complete DENV genomes datasets

DENV-2 whole-genome sequences and associated metadata were obtained from EpiArbo, part of the Global Initiative for Sharing All Influenza Data (GISAID). Sequences were filtered to exclude those with greater than 40% gaps or incomplete sampling dates. The initial dataset was reduced to 7,032 sequences, which then underwent lineage assignment using the most recent dengue lineage implementation in Nextclade^5,49^. Sequences lacking a lineage assignment or displaying poor quality control (QC) scores (based on Nextclade metrics) were discarded.

Final alignments were generated by subsampling within each lineage using Augur^50^. Specifically, one sequence per week per country was selected to ensure balanced temporal and spatial representation, with the exception of Colombia, for which all available sequences were retained. Due tot his sampling asymmetry, trunk reward time estimates for Colombia may be upwardly biased. To detect and remove temporal outliers, a root-to-tip regression was performed. A time-calibrated maximum-likelihood tree was first constructed under a general time-reversible (GTR) model using a fixed molecular clock rate of 8 *×* 10^−4^ substitutions/site/year^51^. Sequences deviating by more than three standard deviations from the regression line were excluded. The tree optimization and temporal calibration were carried out using Augur in conjunction with Treetime^52^.

#### Maximum-likelihood analysis

Maximum-likelihood phylogenetic reconstruction was performed using IQ-tree^53^ under the GTR substitution model^51^. Model selection and branch support were assessed via ModelFinder^54^ and UFBoot^55^ with 1,000 replicates.

#### Discrete phylogeography

Discrete phylogeographic reconstruction was carried out to infer geographic state transitions over time. Prior to phylogeographic inference, time-scaled trees were first generated using BEAST v1.10.5^56^ under a GTR nucleotide substitution model with a molecular clock, excluding discrete traits, with a MCMC chain length of 500 million steps. This preliminary analysis enabled estimation of lineage divergence times and provided an empirical framework for evaluating temporal signal and introduction timing. Using a discrete trait analysis as implemented in BEAST1.10.5^56^, we assigned each sequence a state corresponding to its country of origin. The reconstruction employed Bayesian stochastic search variable^57^ selection to identify the set of well-supported migration events, and state transition probabilities were inferred from the maximum clade credibility (MCC) tree. This analysis enabled the identification of both exportation events from Colombia and introductions into neighboring countries. The resulting posterior trees were analyzed using custom Python scripts to count the number of introductions per year and the inferred timing of viral flow between discrete states.

Introductions were defined as inferred state transitions between discrete geographic locations across the posterior distribution of trees, the timen of each introduction was assigned to the node where the location change occurred in the MCC tree.

#### Poisson Generalized Linear Model of Markov Jumps

To evaluate the role of Colombia in mediating inter-regional Markov Jumps—defined here as discrete state transitions in a phylogeographic reconstruction—we modeled the count of Markov Jumps as a function of multiple covariates using a Poisson generalized linear model (GLM) with a log link function. First, we aggregated the Markov Jump counts by calendar year (or a transformed, centered version of the year variable), by whether the jump involved Colombia, and by the direction of movement (e.g., South to North or North to South). Each aggregated cell thus represented the total number of Markov Jumps meeting the specified criteria within a given year.

Because count data are naturally modeled under a Poisson distribution, and to maintain interpretability in terms of multiplicative changes in the expected number of jumps, we employed the following GLM structure:

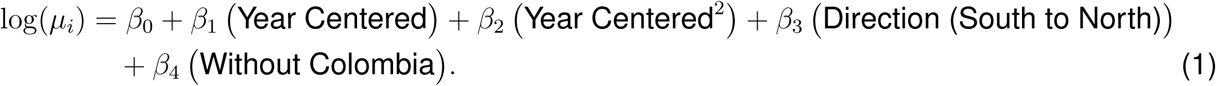

where *µ_i_* denotes the expected count of Markov Jumps in cell *i*. The intercept *β*_0_ represents the baseline log-mean count when all other predictors are at their reference levels (e.g., the reference direction and reference condition “With Colombia”). The “Year Centered” variable adjusts for temporal trends, and a squared term Year Centered^2^ accommodates potential non-linear dynamics. Categorical predictors, such as direction and the Colombia-involvement indicator, were encoded as dummy variables with one category serving as the baseline. Model coefficients *β_k_* were estimated via maximum likelihood, and 95% confidence intervals were computed using the standard errors derived from the Fisher information matrix.

#### Bootstrap-based Source–Sink Score (SSS) analysis

To further evaluate whether the apparent centrality of Colombia in regional viral flow could result from sampling asymmetry, we implemented a complementary bootstrap-based source–sink score (SSS) analysis^28^. The SSS quantifies directional imbalance in phylogeographic transitions, where positive values indicate source-leaning behavior and negative values indicate sink-leaning behavior. We computed SSS values under two independent frameworks: (1) a maximum-likelihood (ML) reconstruction comprising all 1,948 DENV2 II-F.1.1.2 genomes from the Americas, and (2) a time-scaled BEAST reconstruction derived from a temporally and geographically subsampled dataset used in the main analysis.

For each framework, we generated 5,000 bootstrap replicates using three resampling schemes designed to test robustness: (i) *event bootstrap*, which resamples inferred transition events with replacement; (ii) *time-stratified bootstrap*, which preserves the temporal composition of transitions within time bins; and (iii) *tip-jackknife*, which omits a random subset of tips (5% per replicate) before recalculating the transition table. Each resampled dataset yielded a distribution of SSS values from which 95% confidence intervals were computed.

#### Collection, aggregation, and curation of climate and epidemiological data

Climate data was collected from https://www.datos.gov.co/ as a historical record of temperature and humidity as measured by IDEAM’s meterological stations. For the sake of our analyses, only mean temperature and mean relative humidity was used, and assumed to be constant throughout the day. To ensure completeness of data, the metereological conditions of the capital city of each department was used as input for the calculation of the mosquito suitability index. Epidemiological data was collected from Colombia’s National Health Institute (https://portalsivigila.ins.gov.co/) as a historic anonymized dataset that included all suspected and confirmed dengue cases since 2002. The epidemiological data was then filtered to remove records prior to 2012, and was cleaning using custom Python scripts to ensure that cases were assigned to its correct department of reporting. Finally, climate and epidemiological data was aggregated at both department and natural region level for downstream analysis.

#### Estimation of mosquito suitability index (indexP)

The mosquito suitability index was calculated using the previously curated metereological data using the MVSE package in R^58^ using the *Ae. aegypti* prior preset.

#### Dimensionality Reduction via t-Distributed Stochastic Neighbor Embedding (t-SNE)

To visualize the high-dimensional genetic relationships among the envelope protein sequences of 2III D.2, 2II F.1.1.2 and 3III C.2, we first computed a pairwise distance matrix and then embedded these distances in two dimensions using t-SNE in scikit-learn^59^ as follows:

1. Let *n* be the total number of sequences (both serotypes combined). We initialized an *n × n* matrix *D* of zeros.
2. For each pair of sequences (*s_i_, s_j_*), we identified all aligned positions where neither has a gap (“–”). Denote the set of such positions by *M_ij_*.
3. The genetic distance *D_ij_* was then computed as:

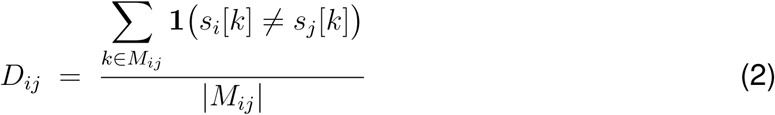

Where **1**(*·*) is the indicator function. This yields the proportion of mismatched bases among non-gapped sites, ensuring that insertions/deletions do not artificially inflate distances.

We then employed scikit-learn’s tSNE class with the precomputed matrix, allowing direct input of the distance matrix D, with two components for two-dimensional visualization. Initialization was set to random to avoid bias from PCA, with a perplexity value of 30 and a random state set to 0 to ensure reproducibility. The resulting t-SNE coordinates were plotted using Matplotlib, with each point colored according to its Nextstrain-assigned clade and connected to an unrooted phylogenetic tree to assess the concordance between tree topology, sequence similarity and the tSNE scatterplot.

#### Additional resources

The detailed protocol for the full-genome sequencing of DENV can be found at https://www. protocols.io/view/dengueseq-a-pan-serotype-whole-genome-amplicon-seq-kqdg39xxeg25/ ^v360^

## Supplemental information index

Figures S1-S4 and their legends in a PDF

Table S1. Accession number and metadata for the sequences generated for this study. Table S2. GISAID Originating laboratories acknowledgement table.

## Data Availability

- All the generated sequences have been deposited and published in GenBank under the accession numbers PQ851425-PQ851490.
- The findings of this study are partially based on metadata and genome sequences associated with 26,633 DENV-2 and 10,753 DENV-3 sequences available on GISAID up to July 26, 2025, and accessible at https://doi.org/10.55876/gis8.250726uw and https://doi.org/10.55876/gis8.250726uh.
- All code required to reproduce the figures and analyses done in this study is available at Github (https://github.com/RicardoRH96/DENV_Epi).
- Any additional information required to reanalyze the data reported in this work paper is available from the lead contact upon request.

https://doi.org/10.55876/gis8.250726uw

https://doi.org/10.55876/gis8.250726uh

## Acknowledgments

We gratefully acknowledge all data contributors, i.e., the Authors and their Originating laboratories responsible for obtaining the specimens, and their Submitting laboratories for generating the genetic sequence and metadata and sharing via the GISAID Initiative, on which this research is partially based.

Ethical approval for the Universidad de Córdoba was granted by the Ethics Committee of the Faculty of Veterinary Medicine and Animal Science, Universidad de Córdoba, together with the Ethics Committee of Hospital San Jerónimo de Montería (meeting minutes No. 011-2021); these activities were carried out under Inter-Institutional Specific Cooperation Agreement No. 018-21 between the two institutions. Independent approval for the sample collection and viral genome sequencing was obtained from the Research Ethics Committee of Hospital Universitario del Valle and the Yale University Human Research Protection Program (protocol No. 2000033281) under their corresponding collaboration agreement. All procedures complied with the Declaration of Helsinki and relevant national regulations. This work was supported by the National Institute of Allergy and Infectious Diseases, National Institutes of Health (award DP2AI176740 to N.D.G.) and by an NIH Shared Equipment grant (1S10OD028669-01) to the Yale Center for Genome Analysis; the funders had no role in study design, data collection and analysis, decision to publish, or manuscript preparation. The findings and conclusions expressed herein are solely those of the authors and do not necessarily represent the official views of the NIH. We thank Clínica Salud Social for generously providing serum samples from dengue-positive patients.

## Author contributions

Conceptualization, R.R, M.S.V, V.T, S.M; methodology, R.R, L.D, V.H, N.D.G, S.M; sample collection and sequencing, R.R, D.P, D.T, D.D, E.L.M, M.I.B, V.T; investigation, R.R, V.T, L.D, D.E.D; writing – original draft, R.R and D.E.D; writing – review & editing, All authors reviewed and approved the manuscript; funding acquisition, N.D.G and S.M; resources, N.D.G and S.M; supervision, N.D.G and S.M.

## Declaration of generative AI and AI-assisted technologies

During the preparation of this work, R.R used OpenAI’s ChatGPT o3 in order to enhance the reusability of the code by helping in function description through comments. After using this tool or service, the author(s) reviewed and edited the content as needed and take(s) full responsibility for the content of the publication.

---

**Supplemental figure 1.**
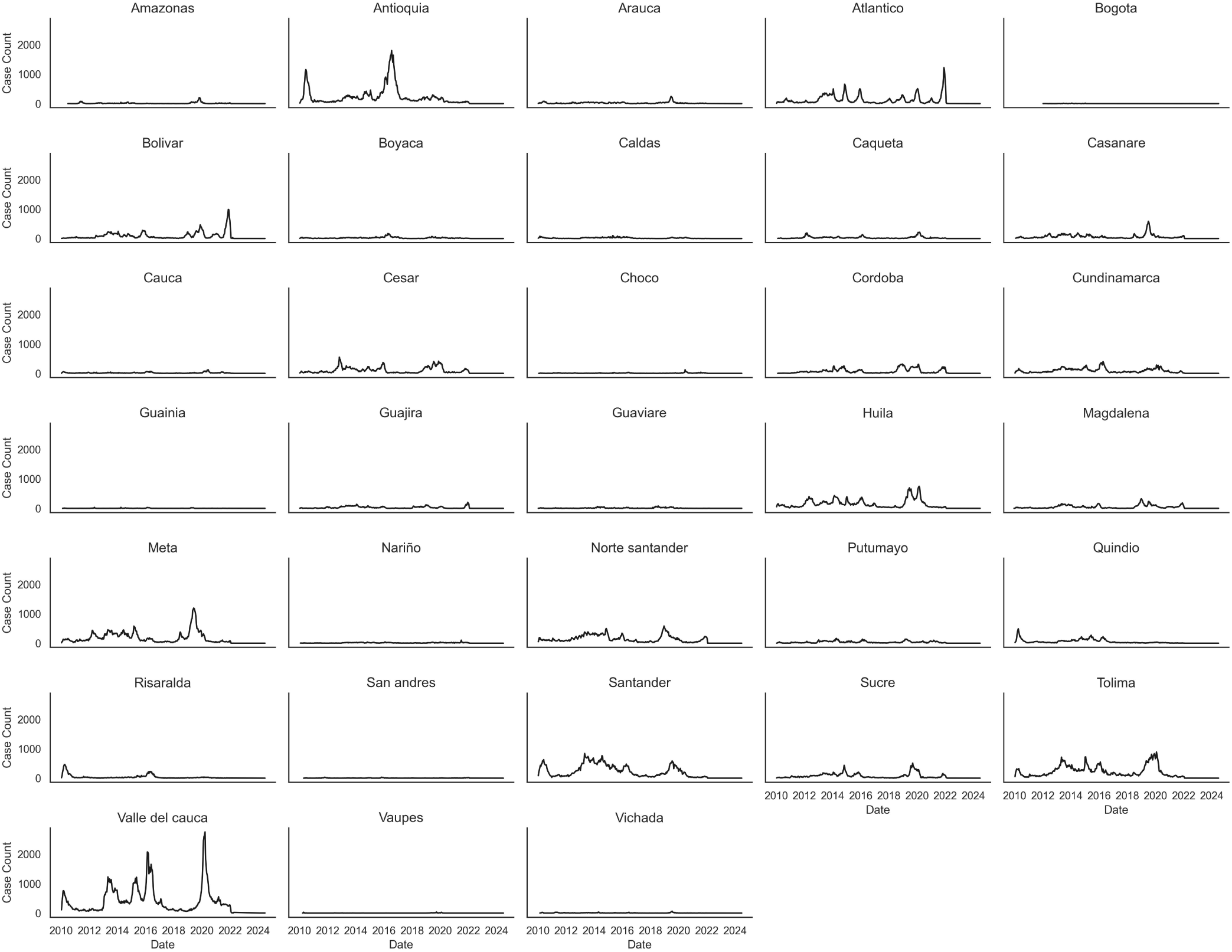
Number of dengue cases for Colombia’s departments.

**Supplemental figure 2.**
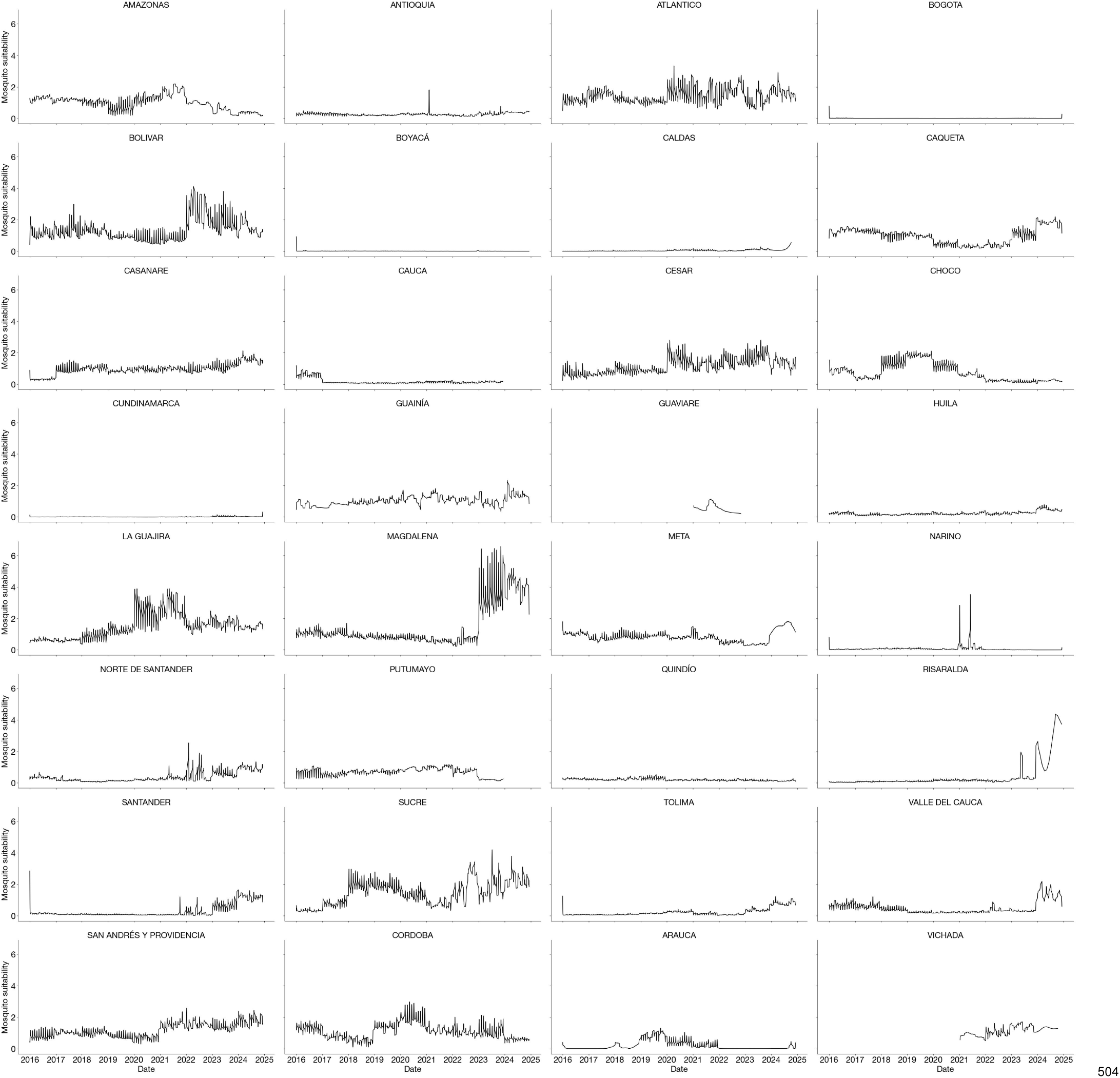
Mosquito suitability index (index P) for Colombian departments.

**Supplemental figure 3.**
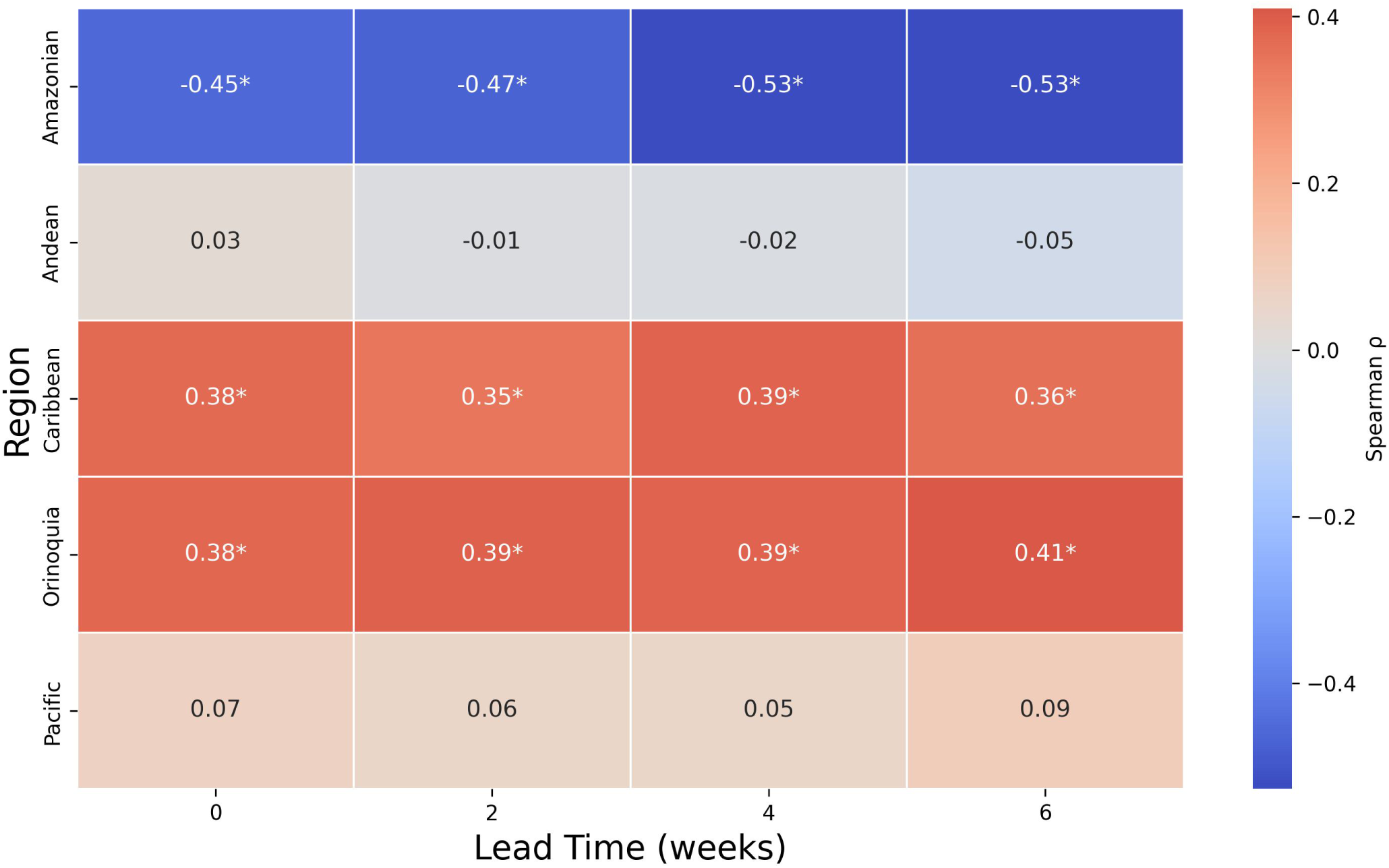
Correlation plot between index P and dengue cases across the natural regions of Colombia at 0-6 weeks of index P lead time. The asterisks next to Spearman’s *ρ* values represent significant correlation (p < 0.05)

**Supplemental figure 4.**
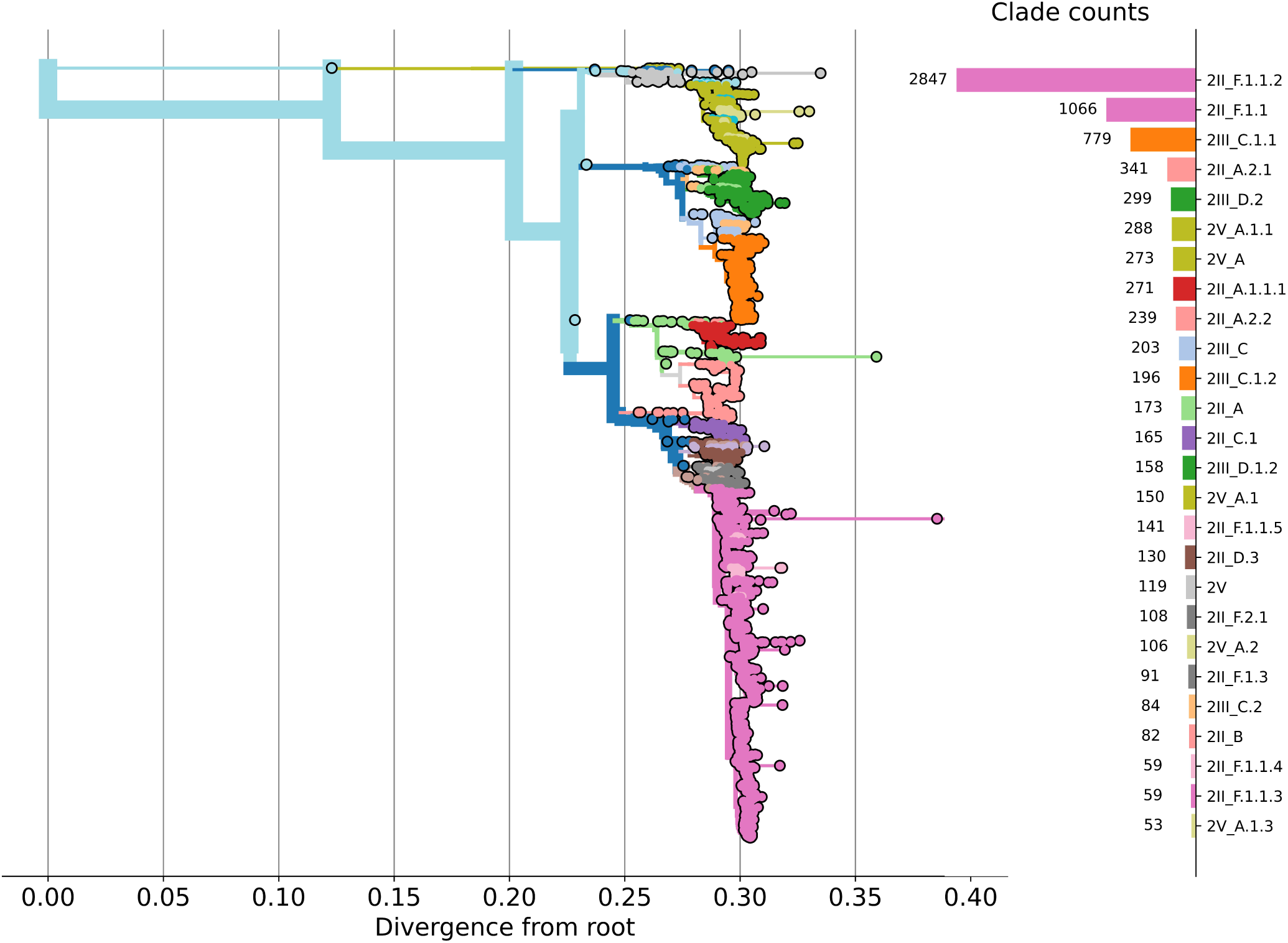
Maximum-likelihood tree of the global diversity of DENV-2 lineages. All the DENV-2 genomes publicly available in Epiarbo were downloaded and processed using the Nextclade CLI to classify them according to its lineage, thickness of the branches represent the number of sequences descending from the branch

**Supplemental figure 5.**
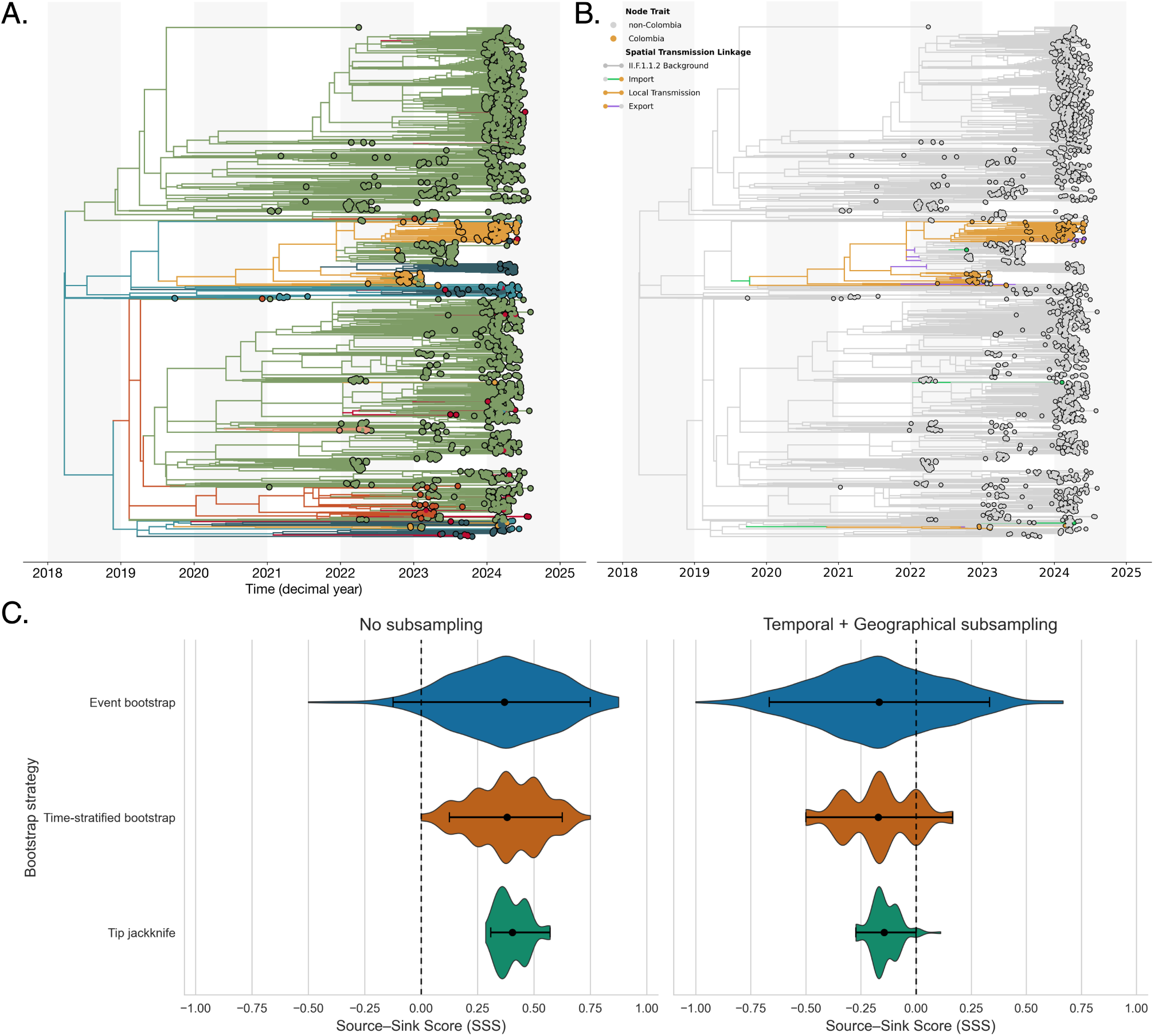
Phylogeographic structure and robustness of 2 IIF.1. lineage dissemination in the Americas. All 1,948 DENV2 II-F.1.1.2 genomes publicly available in EpiArbo were processed using the Nextstrain Dengue build under the mugration model to reconstruct the phylogeographic history of this lineage across the Americas. (A) Maximum-likelihood phylogenetic tree showing Colombian sequences in the context of the continental background, colored by country following the scheme in Figure 3 of the main text. (B) The same tree annotated by Markov-jump transitions through Colombia, with branch colors representing the type of spatial transmission event (importation, local transmission, or export). (C) Bootstrap distributions of the source–sink score (SSS) for Colombia under two independent phylogeographic frameworks: a full maximum-likelihood reconstruction (no subsampling) and a temporally + geographically subsampled BEAST reconstruction. Violin widths represent the density of 5,000 bootstrap replicates for event, time-stratified, and tip-jackknife schemes; dots and horizontal bars denote the mean and 95% CI, and the dashed line marks neutrality (*SSS* = 0). Both frameworks yield small-magnitude, near-neutral SSS values (*|SSS| <* 0.4) with overlapping confidence intervals, indicating that Colombia acts primarily as a hub mediating bidirectional viral exchange rather than as a unidirectional source or sink.

**Supplemental figure 6.**
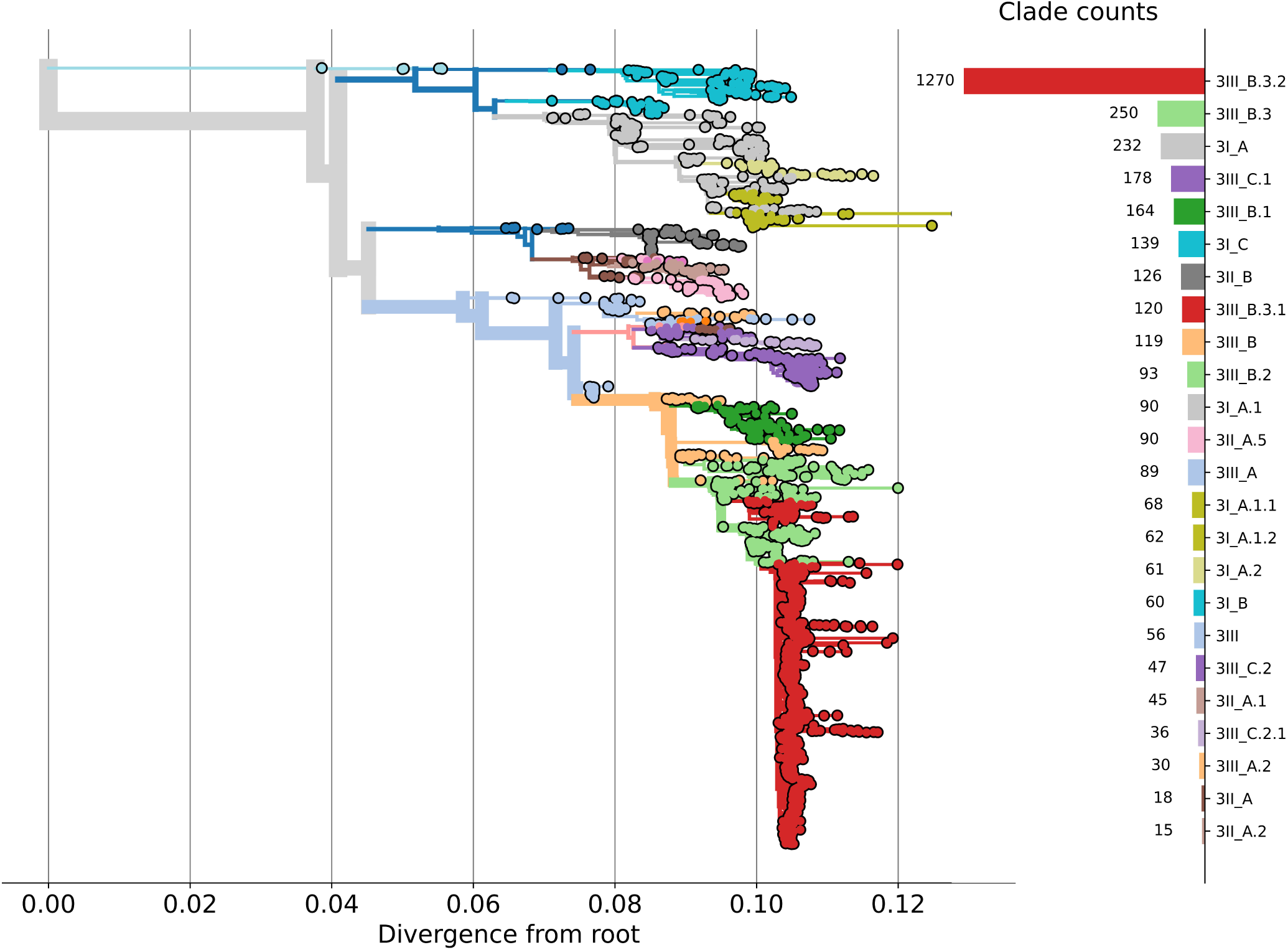
Maximum-likelihood tree of the global diversity of DENV-3 lineages. All the DENV-3 genomes publicly available in Epiarbo were downloaded and processed using the Nextclade CLI to classify them according to its lineage, thickness of the branches represent the number of sequences descending from the branch

**Table 1.** Top 10 enriched biological processes (GO terms) identified by WebGestalt for differentially methylated regions (DMRs) in head tissue, in LPS-treated Cdh1 semi-floxed embryos relative to controls.

| Study-ID | PCR-CT | Serotype | Lineage | Genome Coverage | Collection Date | City | GenBank-ID |
| --- | --- | --- | --- | --- | --- | --- | --- |
| D0348 | 26.95 | DENV2 | 2II.F.1.1.2 | 99.07 | 2024-03-13 | Cali | PQ851425 |
| D0350 | 30.15 | DENV2 | 2II.F.1.1.2 | 91.33 | 2024-03-15 | Vila Rica, Cauca | PQ851426 |
| D0353 | 15.24 | DENV2 | 2III.D.2 | 100.0 | 2024-03-15 | Cali | † |
| D0360 | 21.06 | DENV2 | 2III.D.2 | 100.0 | 2024-03-20 | Candelaria | PQ851427 |
| D0368 | 25.24 | DENV2 | 2III.D.2 | 98.0 | 2024-03-21 | Cali | PQ851428 |
| D0370 | 31.13 | DENV2 | 2III.D.2 | 91.38 | 2024-03-26 | Cali | PQ851429 |
| D0372 | 19.96 | DENV2 | 2III.D.2 | 100.0 | 2024-03-25 | Cali | PQ851430 |
| D0373 | 18.02 | DENV2 | 2III.D.2 | 100.0 | 2024-03-26 | Corinto | PQ851431 |
| D0380 | 26.0 | DENV2 | 2II.F.1.1.2 | 100.0 | 2024-04-02 | Santander De Quilichao | PQ851432 |
| D0381 | 11.47 | DENV2 | 2II.F.1.1.2 | 100.0 | 2024-04-01 | Pradera | PQ851433 |
| D0384 | 35.01 | DENV2 | 2II.F.1.1.2 | 68.54 | 2024-04-02 | Cali | PQ851434 |
| D0385 | 34.19 | DENV2 | 2III.D.2 | 85.13 | 2024-04-03 | Cali | PQ851435 |
| D0389 | 28.46 | DENV2 | 2II.F.1.1.2 | 92.47 | 2024-04-08 | Yumbo | PQ851436 |
| D0394 | 27.4 | DENV2 | 2II.F.1.1.2 | 99.6 | 2024-04-06 | Cali | PQ851437 |
| D0397 | 28.27 | DENV2 | 2III.D.2 | 94.35 | 2024-04-08 | Cali | PQ851438 |
| D0399 | 27.44 | DENV2 | 2II.F.1.1.5 | 99.35 | 2024-04-10 | Cali | PQ851439 |
| D0401 | 20.82 | DENV2 | 2III.D.2 | 100.0 | 2024-04-11 | Cali | PQ851440 |
| D0403 | 34.13 | DENV2 | 2II.F.1.1.2 | 77.01 | 2024-04-13 | Palmira | PQ851441 |
| D0404 | 16.75 | DENV2 | 2II.F.1.1.2 | 100.0 | 2024-04-11 | Cali | PQ851442 |
| D0409 | 24.26 | DENV2 | 2II.F.1.1.2 | 100.0 | 2024-04-18 | Cali | PQ851443 |
| D0413 | 35.22 | DENV2 | 2II.F.1.1.2 | 64.85 | 2024-04-17 | Cali | * |
| D0414 | 31.4 | DENV2 | 2II.F.1.1.2 | 91.38 | 2024-04-21 | Cali | PQ851444 |
| D0415 | 29.04 | DENV2 | 2III.D.2 | 98.95 | 2024-04-19 | Cali | PQ851445 |
| D0417 | 20.24 | DENV2 | 2II.F.1.1.2 | 100.0 | 2024-04-21 | Cali | PQ851446 |
| D0421 | 35.87 | DENV2 | 2II.F.1.1.2 | 63.97 | 2024-04-22 | Cali | * |
| D0425 | 17.16 | DENV2 | 2III.D.2 | 100.0 | 2024-04-24 | Cali | PQ851447 |
| D0427 | 33.69 | DENV2 | 2II.F.1.1.2 | 82.32 | 2024-04-25 | Mariano Ramos | PQ851448 |
| D0430 | 34.62 | DENV2 | 2II.F.1.1.2 | 85.09 | 2024-04-27 | Rozo, Valle | PQ851449 |
| D0434 | 34.61 | DENV2 | 2II.F.1.1.2 | 57.33 | 2024-05-05 | Bella Vista | * |
| D0435 | 30.69 | DENV2 | 2II.F.1.1.2 | 80.45 | 2024-05-07 | Cali | PQ851450 |
| D0439 | 28.1 | DENV2 | 2II.F.1.1.2 | 96.41 | 2024-05-08 | Cali | PQ851451 |
| D0440 | 28.06 | DENV2 | 2II.F.1.1.2 | 98.04 | 2024-05-08 | Cali | PQ851452 |
| D0443 | 27.88 | DENV2 | 2II.F.1.1.2 | 97.84 | 2024-05-07 | Cali | PQ851453 |
| D0446 | 22.29 | DENV2 | 2II.F.1.1.2 | 99.98 | 2024-05-12 | Cali | PQ851454 |
| D0451 | 34.97 | DENV2 | 2II.F.1.1.2 | 63.35 | 2024-05-13 | Cali | * |
| D0456 | 29.17 | DENV2 | 2III.D.2 | 97.35 | 2024-05-15 | Corinto | † |
| D0461 | 29.18 | DENV2 | 2III.D.2 | 90.98 | 2024-05-18 | Santander De Quilichao | PQ851455 |
| D0463 | 29.23 | DENV2 | 2II.F.1.1.2 | 94.62 | 2024-05-18 | Cali | PQ851456 |
| D0464 | 15.39 | DENV2 | 2II.F.1.1.2 | 100.0 | 2024-05-19 | Cali | PQ851457 |
| D0465 | 23.38 | DENV2 | 2II.F.1.1.2 | 99.98 | 2024-05-18 | Cali | PQ851458 |
| D0467 | 32.99 | DENV2 | 2II.F.1.1.2 | 87.53 | 2024-05-23 | Yotoco | PQ851459 |
| D0471 | 23.64 | DENV2 | 2II.F.1.1.2 | 99.98 | 2024-05-25 | Cali | PQ851460 |
| D0473 | 26.89 | DENV2 | 2II.F.1.1.2 | 97.05 | 2024-05-26 | Cali | PQ851461 |
| D0480 | 31.59 | DENV2 | 2III.D.2 | 84.52 | 2024-05-30 | Cali | PQ851462 |
| D0489 | 28.56 | DENV2 | 2II.F.1.1.2 | 92.94 | 2024-06-06 | Cali | PQ851463 |
| D0491 | 26.97 | DENV2 | 2II.F.1.1.2 | 99.59 | 2024-06-07 | Palmira | PQ851464 |
| D0495 | 27.36 | DENV2 | 2II.F.1.1.2 | 98.6 | 2024-06-11 | Cali | PQ851465 |
| D0498 | 32.55 | DENV2 | 2III.D.2 | 76.06 | 2024-06-10 | Cali | PQ851466 |
| D0499 | 32.83 | DENV2 | 2II.F.1.1.2 | 71.8 | 2024-06-09 | Vijes | PQ851467 |
| D0523 | 21.94 | DENV2 | 2II.F.1.1.2 | 97.05 | 2024-06-20 | Palmira | PQ851468 |
| D0524 | 21.62 | DENV2 | 2II.F.1.1.2 | 98.52 | 2024-06-22 | Cali | PQ851469 |
| D0525 | 20.46 | DENV2 | 2II.F.1.1.2 | 99.58 | 2024-06-23 | Cali | PQ851470 |
| D0526 | 17.27 | DENV2 | 2II.F.1.1.2 | 99.78 | 2024-06-23 | Cali | PQ851471 |
| D0533 | 19.34 | DENV2 | 2II.F.1.1.2 | 100.0 | 2024-06-27 | Jamundi | PQ851472 |
| D0537 | 31.09 | DENV2 | 2II.F.1.1.2 | 81.02 | 2024-07-02 | Cali | PQ851473 |
| D0540 | 25.03 | DENV2 | 2II.F.1.1.2 | 95.6 | 2024-07-05 | Candelaria | PQ851474 |
| D0544 | 28.31 | DENV2 | 2II.F.1.1.2 | 87.77 | 2024-07-13 | Cali | PQ851475 |
| D0550 | 20.83 | DENV2 | 2II.F.1.1.2 | 99.78 | 2024-07-20 | Cali | PQ851476 |
| D0560 | 30.38 | DENV2 | 2II.F.1.1.2 | 87.69 | 2024-07-26 | Cali | PQ851477 |
| D0566 | 17.59 | DENV2 | 2II.F.1.1.2 | 100.0 | 2024-07-31 | Cali | PQ851478 |
| D0567 | 17.88 | DENV2 | 2II.F.1.1.2 | 100.0 | 2024-07-28 | Cali | PQ851479 |
| D0568 | 32.74 | DENV2 | 2II.F.1.1.2 | 72.44 | 2024-08-01 | Cali | PQ851480 |
| D0569 | 26.87 | DENV2 | 2II.F.1.1.2 | 93.45 | 2024-08-01 | Cali | PQ851481 |
| D0573 | 31.1 | DENV2 | 2II.F.1.1.2 | 82.25 | 2024-08-03 | Caloto, Cauca | PQ851482 |
| D0575 |  | DENV2 | 2II.F.1.1.2 | 85.86 | 2024-08-07 | Putumayo | PQ851483 |
| D0576 | 34.43 | DENV2 | 2II.F.1.1.2 | 86.45 | 2024-08-07 | Santander De Quilichao | PQ851484 |
| D0577 | 26.24 | DENV2 | 2II.F.1.1.5 | 98.61 | 2024-08-06 | Cali | PQ851485 |
| D0578 | 25.33 | DENV2 | 2II.F.1.1.2 | 92.89 | 2024-08-07 | Cali | PQ851486 |
| D0476 | 28.5 | DENV2 | 2II.F.1.1.2 | 95.93 | 2024-05-28 | Cali | PQ851487 |
| D0478 | 24.2 | DENV2 | 2II.F.1.1.2 | 100.0 | 2024-05-28 | Cali | PQ851488 |
| D0585 | 33.19 | DENV2 | 2II.F.1.1.2 | 65.44 | 2024-08-14 | Cali | * |
| D0586 | 33.3 | DENV2 | 2II.F.1.1.2 | 86.41 | 2024-08-15 | Cali | PQ851489 |
| D0587 | 31.47 | DENV2 | 2II.F.1.1.2 | 89.35 | 2024-08-08 | Cali | PQ851490 |

**Notes:**
\* %Coverage below the threshold for submission; † Suspected co-infection or cross-contamination

